# Infectivity of symptomatic malaria patients to *Anopheles farauti* colony mosquitoes in Papua New Guinea

**DOI:** 10.1101/2020.06.02.20120816

**Authors:** Lincoln Timinao, Rebecca Vinit, Michelle Katusele, Tamarah Koleala, Elma Nate, Cyrille Czeher, Louis Schofield, Ingrid Felger, Ivo Mueller, Moses Laman, Leanne J. Robinson, Stephan Karl

## Abstract

Despite being a weak point in their life cycle, transmission of *Plasmodium* parasites from humans to mosquitoes is an understudied field of research. Direct membrane feeding assays (DMFA) are an important tool, allowing detailed mechanistic malaria transmission studies from humans to mosquitoes. Especially for *Plasmodium vivax*, which cannot be cultured long-term under laboratory conditions, implementation of DMFAs requires proximity to *P. vivax* endemic areas. In the present study, we investigated the infectivity of symptomatic *Plasmodium* infections to *Anopheles farauti* colony mosquitoes in Papua New Guinea (PNG), a country with one of the highest rates of *Plasmodium vivax* in the world. A total of 182 DMFAs were performed with venous blood collected from symptomatic malaria patients positive by rapid diagnostic test (RDT). DMFAs resulted in mosquito infection in 20.9% (38/182) of cases. The parasite species in the blood feeds were determined retrospectively by expert light microscopy and quantitative real-time qPCR. Based on light microscopy, 9.2% of *P. falciparum* and 42% of *P. vivax* human infections resulted in mosquito infections. Infections containing gametocytes detected by microscopy led to mosquito infections in 58.8% of *P. vivax* and 8.7% of *P. falciparum* infections. Based on qPCR, 10% of *P. falciparum* and 43.6% of *P. vivax* lead to a successful mosquito infection. Venous blood samples from symptomatic *P. vivax* patients were more infectious to *An. farauti* mosquitoes in DMFAs compared to *P. falciparum* infected patients. The capacity to perform DMFAs in a high-burden *P. vivax* setting creates a unique opportunity to address critical gaps in our understanding of *P. vivax* human-tomosquito transmission.

## 1. Introduction

Transmission between the human host and the mosquito vector is a crucial step in the malaria parasite life cycle. It represents a bottleneck where parasite numbers shrink from millions in the human body to less than a hundred in the mosquito vector. (Smith et al., 2014) Transmission through the mosquito thus represents a vulnerable target, with the potential to be interrupted, if methods can be devised to prevent parasites from propagating in the mosquito and reaching the sporozoite stage (Sauerwein and Bousema, 2015). Studying human to mosquito transmission in order to identify potential pathways to prevent or block transmission is a key focus of malaria research (Churcher et al., 2015; Sauerwein and Bousema, 2015) and as such, research tools have been designed to explore this transitioning phase of the parasite. Direct membrane feeding assays (DMFAs) are one such tool. DMFAs were initially developed by Rutledge and colleagues in 1964 and it involve artificially exposing malaria parasites to mosquitoes via a membrane feeding apparatus (Rutledge et al., 1964).

DMFAs provide a means to investigate the still poorly understood process of human to mosquito transmission and the resulting mosquito infection in a controlled research environment. For example, DMFAs can be used to study the infectiousness of different human malaria reservoirs, and estimate their contribution towards transmission (Diallo et al., 2008; Graves et al., 1988). This can include symptomatic, patent infections as in the present study and asymptomatic, often low-density infections. (Kiattibutr et al., 2017) In addition, DMFAs can be used to study the effect of drugs, vaccine candidates and immune factors on the development of the mosquito stages of the Plasmodium parasites (Bousema et al., 2012; Delves et al., 2012; Sattabongkot et al., 2015; Vallejo et al., 2016). Finally, DMFAs provide an opportunity for circumventing some of the operational and ethical complicating factors associated with feeding mosquitoes directly on the skin of malaria infected individuals.

Despite these advantages, DMFAs are resource intensive, require an insectary and rely on stringent logistics for sample collection, handling, rapid transportation and processing as it has been shown that the time between blood collection and performance of the DMFA can impact assay outcome, most likely due to premature gametocyte activation (Churcher et al., 2012; Sattabongkot et al., 2015). As a further complication, conducting DMFAs with *P. vivax* requires proximity to endemic areas in order to access infected samples as continuous culture of this parasite species remains elusive (Roobsoong et al., 2015). Papua New Guinea (PNG) has the highest *P. vivax* burden in the world, thus *P. vivax* is a research priority for the country and infected blood samples can still easily be obtained (Cattani et al., 1986; Howes et al., 2016; Müller et al., 2003; World Health Organization, 2019). Establishing DMFAs with *P. vivax* provides a tool to study *P. vivax* transmission that is of potentially global relevance.

DMFAs were performed in PNG previously in 1983–1985 in village-based malaria surveys, prior to diagnosis and on known gametocyte carriers in clinical outpatient populations in Madang and Goroka (Graves et al., 1988). In the present study, we investigated the infectivity of blood samples obtained from symptomatic, rapid diagnostic test (RDT)-positive individuals to *Anopheles farauti* colony mosquitoes.

## 2. Materials and methods

### 2.1. Sample collection

This study was conducted at the PNG Institute of Medical Research (PNGIMR) in Madang Province, PNG, between May 2014 and November 2018. Study participants were recruited from Madang Town Clinic and Yagaum Rural Health Centre. Ethical approval was received from the PNGIMR Institutional Review Board (IRB #1516) and the PNG Medical Research Advisory Committee (MRAC #16.01). Written informed consent was received from all individuals enrolled in the study. Individuals presenting with malaria symptoms were tested with rapid diagnostic tests (RDT). The current malaria RDTs are immunochromatographic tests that detect the presence of circulating parasite antigens. The two most commonly used antigens are the histidine rich protein-2 (HRP2) which is a protein molecule produced by only *P. falciparum*, hence the detection of this antigen confirms *P. falciparum* infections. The second antigen is the Plasmodium lactate dehydrogenase (pLDH) which is an intracellular metabolic enzyme, present in all *Plasmodium* species and the detection of this antigen would mean infection with any *Plasmodium* species. In the present study, CareStart Malaria Pf/PAN (HRP2/pLDH) Ag Combo RDT (Access Bio, Somerset, NJ, United States) RDTs were used. From RDT-positive individuals venous blood samples (3–5 mL) were collected in Vacutainers (BD, North Ryde, NSW, Australia) and immediately stored in a thermal flask filled with water (∼38.0 °C, measured by a digital thermometer attached to the flask). The blood was transported to the insectary immediately and *An. farauti* colony mosquitoes, which had been starved for 4–5 h, were exposed to blood samples via a water-jacketed membrane feeding apparatus. Time between sampling and feeding is an important parameter. In the present study, the time between sampling and feeding was approximately 20–30 min for samples collected at Yagaum Hospital, located in a 10 min walking distance from the insectary. Transport of blood samples collected in Madang Town Clinic took about 2 h and involved a 30 min drive.

### 2.2. Mosquito colony maintenance, membrane feeding assays and mosquito dissection

The present study used an *An. farauti* colony, which was first adapted in Rabaul, East New Britain province of PNG in 1968. Later in the 1980s males were brought from Rabaul and mated with females from Agan village, Madang province and since then they were used in several studies (Beebe et al., 2000; Collins et al., 2002; Graves et al., 1988; Sweeney, 1987). The initial colony line (obtained from Queensland Institute of Medical Research, Australia in 2009) was backcrossed with wild *An. farauti* mosquitoes at PNGIMR in 2009. However, the genetic contribution of the wild-type backcross to the colony has not been quantified. The colony was maintained using established methods (Nace et al., 2004). To conduct DMFAs, 3–5 day old female mosquitoes from the *An. farauti* colony were separated into paper cups with 50–100 individuals per cup. Each human blood sample was exposed to 100–400 mosquitoes(i.e., 2–4 cups with 50–100 mosquitoes).

The feeding cups and apparatus were set up in the laboratory prior to the arrival of the samples in order to minimise the time between sample collection and feeding. The light in the insectary was dimmed and the apparatus was covered with a dark cloth for the period of feeding. After allowing the mosquitoes to feed for ∼15–20 min, any unfed mosquitoes were removed. The cups containing the fed mosquitoes were kept for 7–9 days and were dissected for *P. vivax* and *P. falciparum* oocysts (Ouedraogo et al., 2013; Sattabongkot et al., 2015). Between 50 –100 mosquitoes were dissected per DMFA following established methodology (Ouédraogo et al., 2013). Mosquito midguts were stained with 0.2% mercurochrome for 10–15 min and oocysts were counted under a light microscope at 40x magnification.

### 2.3 Light microscopy and PCR detection of malaria parasites

Thick and thin blood films were prepared using standard methodology and were stained with

4% Giemsa stain for 30 min. Slides were read according to WHO standards and by WHO certified microscopists. Parasite density was calculated as the geometric mean of densities obtained from reads by two expert microscopists. If the two reads were discrepant in terms of the presence or absence of parasites, parasite density (i.e. if they differed by a factor of 10) and parasite species the slide was read by a third expert microscopist to resolve the discrepancy. DNA extraction was performed on 250µL of whole blood and a quantitative real-time PCR (qPCR) was performed to quantify the infection and determined the parasite species as described elsewhere. (Wampfler et al., 2013)

### 2.4. Statistical analyses

Prism 6.01 (GraphPad Software, La Jolla, CA USA) and Stata 13 (StataCorp, College Station, TX, USA) were used to analyse data. To compare proportions, two-sample tests of proportions were used. To test the influence of a continuous variable (such as parasite density) on a binary outcome variable (such as DMFA success rate), logistic regression was used. To test the association between two continuous variables such as infection rate in the successfully infected mosquitoes versus gametocyte density we used non-parametric correlation analysis (Spearman’s rank correlation) with a significance level of p = 0.05.

## 3. Results

### 3.1. Study population

Selection of patients relied on RDT diagnosis. Subsequent light microscopy examinations of the corresponding blood slides and molecular diagnosis by qPCR were conducted for 182 RDT-positive participants. Table 1 shows the characteristics of the study population and the results from light microscopy examination and molecular diagnosis by qPCR.

**Table 1:**
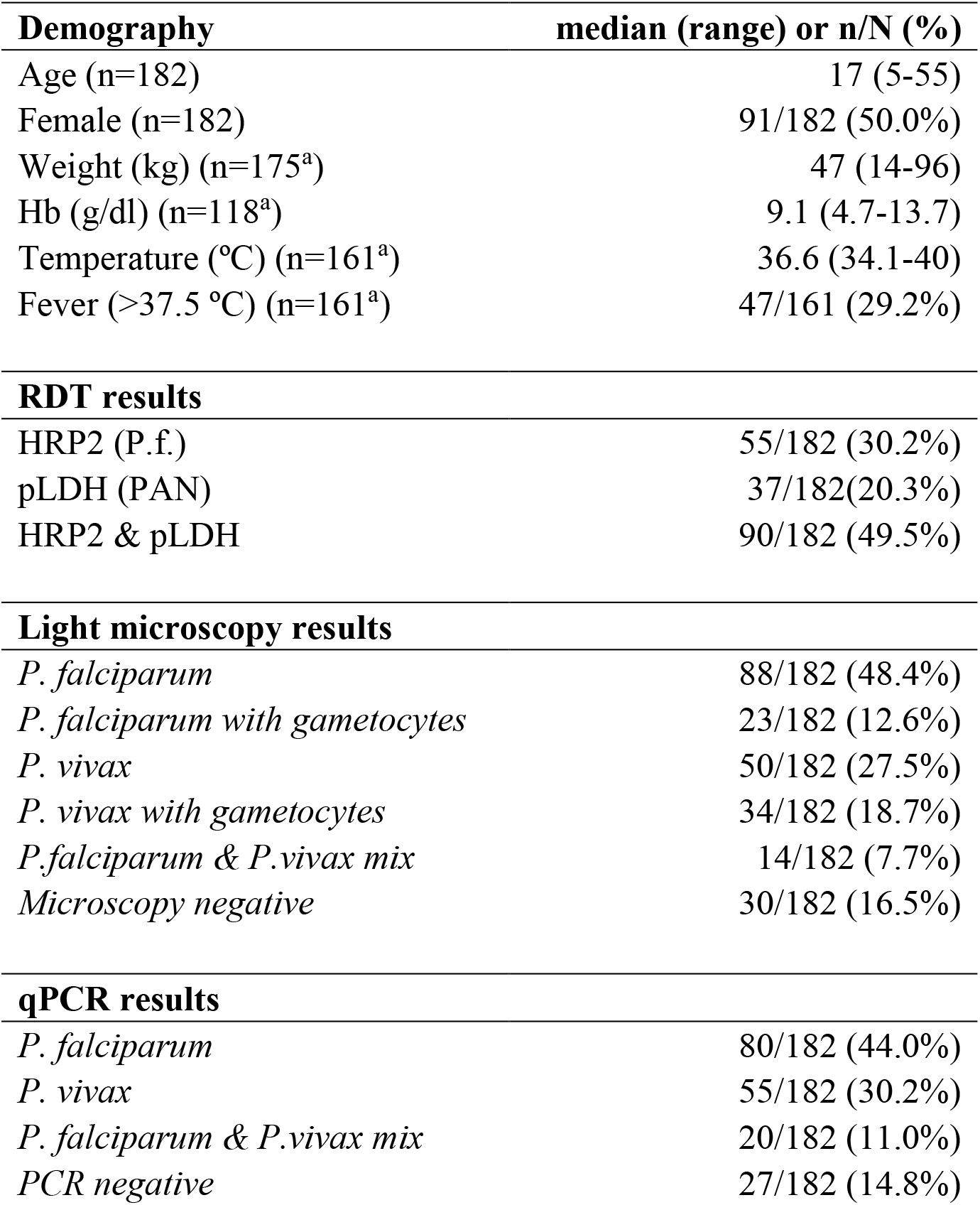
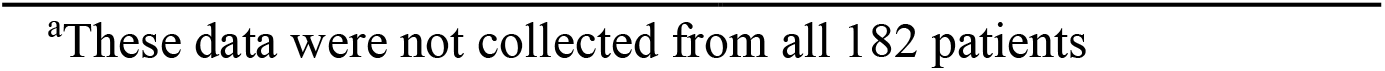
Characteristics of the study population and malaria diagnosis through RDT, microscopy and qPCR. Values are presented as proportions (n/N) and percentage or median and range.

| **Demography** | **median (range) or n/N (%)** |
| --- | --- |
| Age (n=182) | 17 (5-55) |
| Female (n=182) | 91/182 (50.0%) |
| Weight (kg) (n=175<sup>a</sup>) | 47 (14-96) |
| Hb (g/dl) (n=118<sup>a</sup>) | 9.1 (4.7-13.7) |
| Temperature (°C) (n=161<sup>a</sup>) | 36.6 (34.1-40) |
| Fever (>37.5 °C) (n=161<sup>a</sup>) | 47/161 (29.2%) |
| **RDT results** |  |
| HRP2 (P.f.) | 55/182 (30.2%) |
| pLDH (PAN) | 37/182(20.3%) |
| HRP2 & pLDH | 90/182 (49.5%) |
| **Light microscopy results** |  |
| *P. falciparum* | 88/182 (48.4%) |
| *P. falciparum* with gametocytes | 23/182 (12.6%) |
| *P. vivax* | 50/182 (27.5%) |
| *P. vivax* with gametocytes | 34/182 (18.7%) |
| *P.falciparum* & *P.vivax* mix | 14/182 (7.7%) |
| Microscopy negative | 30/182 (16.5%) |
| **qPCR results** |  |
| *P. falciparum* | 80/182 (44.0%) |
| *P. vivax* | 55/182 (30.2%) |
| *P. falciparum* & *P.vivax* mix | 20/182 (11.0%) |
| PCR negative | 27/182 (14.8%) |

<sup>a</sup>These data were not collected from all 182 patients

The largest proportion of individuals (48.9%) was RDT positive for both, HRP2 and pLDH tests while 30.2% and 20.3% of patients were positive only for HRP2 or pLDH-based tests, respectively. Light microscopy revealed that the largest proportion of symptomatic patients in this study population were infected with *P. falciparum* (48.4%) followed by *P. vivax* (27.5%). Median (range) parasite density was 5,600 (8 – 96,000) for *P. falciparum* and 3,744 (8 – 28,109) for *P. vivax*. There were 14 mixed infections (7.7%) containing both, *P. falciparum* and *P. vivax*. The qPCR results revealed a slightly higher proportion of *P. falciparum* infections (44%) than *P. vivax* infections (30.2%). A higher proportion of the samples were diagnosed as mixed infections by qPCR as compared to microscopy (11.1% vs 7.7%). There were 17/182 (9.3%) samples that were negative according to both microscopy and qPCR while 142/182 (78%) were positive by both microscopy and qPCR with a concordance of 87.4%. We observed that 10/182 (5.5%) of samples were negative by qPCR but positive by microscopy while 13/182 (7.1%) of samples were positive by qPCR but negative by microscopy.

### 3.2. DMFA results

Overall, 38/182 (20.9%) DMFAs gave rise to an infection in the mosquitoes. Figure 1 shows an example of an *An. farauti* midgut infected with *P. vivax* oocysts 7 days post infection.

**Figure 1:**
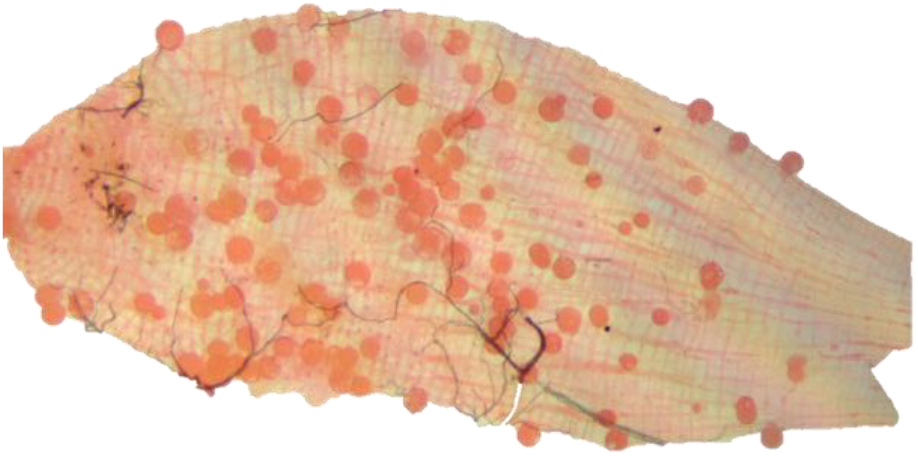
Infected midgut from *An. farauti* mosquito dissected in the present study. The image was taken on a Zeiss Primostar microscope equipped with an Axiocam 105 Color camera (Carl Zeiss Pty. Ltd.) at 40x magnification. The image was then edited using PowerPoint, Microsoft office 2010.

Table 2 shows the proportion of DMFAs leading to an infection in the mosquitoes according to the type of diagnostic result, together with the corresponding average and range of oocysts per midgut. Light microscopy diagnosis revealed, that *P. vivax* infections were more often infectious to the mosquitoes as compared to *P. falciparum* infections (42% vs. 9.2%, p< 0.001). Within the *P. vivax* samples, a higher proportion was infectious to mosquitoes if samples contained detectable numbers of *P. vivax* gametocytes by microscopy (58.8%). We noted that 6.3% (1/16) and 10.9% (7/64) of the *P. vivax* and *P. falciparum* infections that gave rise to infected mosquitoes had no gametocytes by microscopy. In addition, 8 of the 14 (57.1%) mixed infections by microscopy gave rise to mosquito infections.

**Table 2:** DMFAs giving rise to mosquito infections according to RDT, microscopy and qPCR. All samples were collected from symptomatic RDT positive patients. Values are presented either as proportion (n/N) and percent, or as average and minimum to maximum range.

| Diagnostic result | Proportion of DMFAs resulting in mosquito infection; n/N (%) | Oocyst number (average, range)* |
| --- | --- | --- |
| **RDT** |  |  |
| HRP2 | 15/55 (27.3%)<sup>a</sup> | 8 (1-106) |
| pLDH | 13/37 (35.1%)<sup>b</sup> | 36 (1-534) |
| HRP2 + pLDH (both lines) | 10/90 (11.1%)<sup>c</sup> | 4 (1-17) |
| Overall | 38/182 (20.9%) | 15 (1-534) |
| **Light microscopy** |  |  |
| *P. falciparum* | 8/87 (9.2%)<sup>d</sup> | 5 (1-16) |
| *P. falciparum* with gametocytes | 2/23 (8.7%) | 1 (1-2) |
| *P. vivax* | 21/50 (42%)<sup>e</sup> | 17 (1-534) |
| *P. vivax* with gametocytes | 20/34 (58.8%)<sup>f</sup> | 18 (1-534) |
| *P. falciparum* & *P. vivax* mixed | 8/14 (57.1%) | 6 (1-36) |
| Microscopy negative | 1/30 (3.3%) | 13 (1-24) |
| Overall | 38/182 (20.9%) | 15 (1-534) |
| **qPCR** |  |  |
| *P. falciparum* | 8/80 (10.0%) | 5 (1-43) |
| *P. vivax* | 24/55 (43.6%) | 17 (1-534) |
| *P. falciparum* & *P. vivax* mixed | 4/20 (20.0%) | 14 (1-106) |
| PCR negative | 2/27 (7.4%) | 9 (1-24) |
| Overall | 38/182 (20.9%) | 15 (1-534) |

**Note:** \*only infected mosquitoes were considered (i.e., uninfected mosquitoes were not included into this calculation); significant differences were observed in the proportions *a* vs. *b*, *a* vs. *c*; *b* vs. *c*, *d* vs. *e* and *d* vs. *f*.

Infection success, i.e., DMFAs resulting in at least 1 infected mosquito, was not significantly correlated with parasite or gametocyte density when tested using logistic regression in any of these groups (*P. vivax, P. falciparum, P vivax* with gametocytes). The proportion of infected mosquitoes per DMFA and oocyst numbers per midgut were not correlated to *P. falciparum* or *P. vivax* asexual parasite density. Furthermore, there was no correlation between the proportion of infected mosquitoes and the copy numbers of *P. vivax* asexual stages by qPCR.

The proportion of infected mosquitoes was significantly correlated with *P. vivax* gametocyte density (p = 0.049, Spearman’s rank correlation) as shown in Figure 2A. However, the considerable scatter and correlation coefficient of R = 0.42 indicated that the correlation is not very strong. The number of oocysts per dissected mosquito midgut was significantly correlated with the proportion of infected mosquitoes per DMFA (p< 0.0001, Spearman’s rank correlation coefficient R = 0.82) as shown in Figure 2B.

**Figure 2:**
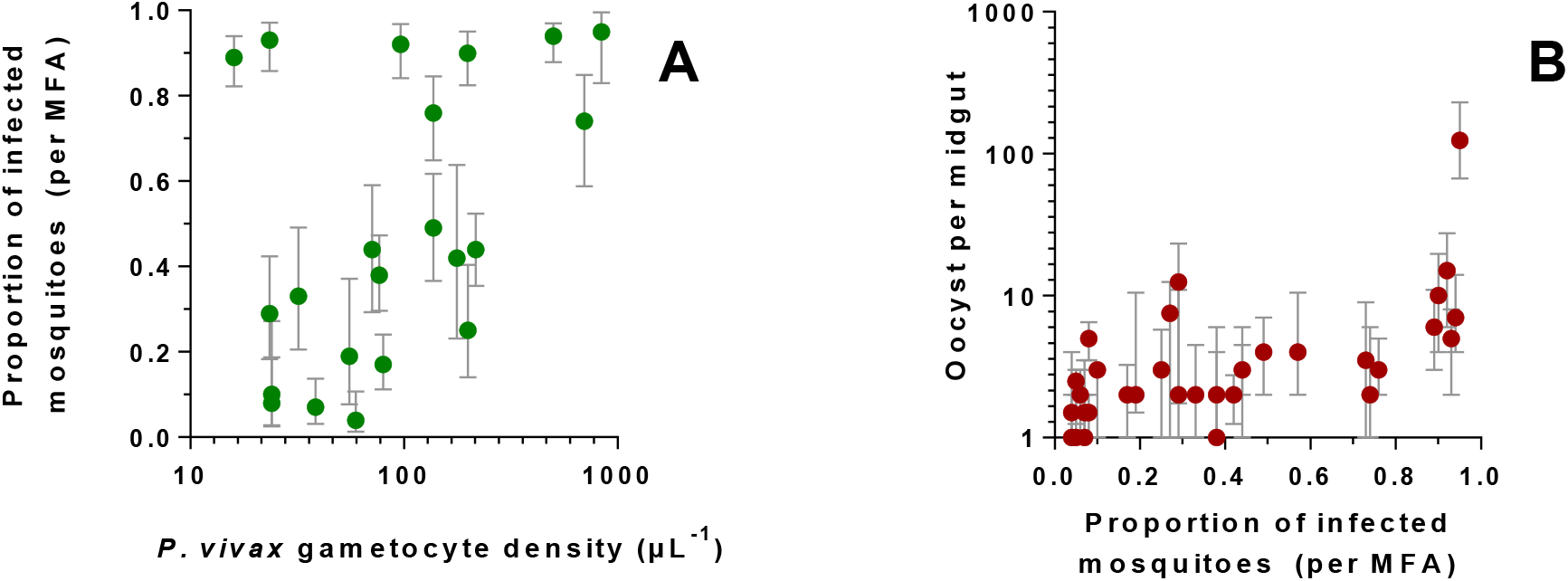
Correlation between mosquito infection success and *P. vivax* gametocyte density or oocyst count. Panel A shows the correlation of the proportion of infected mosquitos per DMFA with *P. vivax* gametocyte density. The error bars are 95% confidence intervals of proportions. Panel B shows the correlation between the proportion of infected mosquitoes per DMFA and the number of oocysts per infected midgut. The error bars are interquartile ranges.

## 4. Discussion

DMFAs with *P. falciparum* can be conducted in well-established laboratories worldwide, including non-endemic settings, due to the straight-forward and well-established *in vitro* culture method available for this species. This is currently not the case with *P. vivax* due to the limitations with continuous parasite culture necessitating access to naturally acquired infections in field settings, often associated with additional cost and operational constrains. As such, a reliable *P. vivax* DMFA set up is of great value.

In the present study, we established such a setup in PNG and investigated the infectiousness of symptomatic, RDT-positive malaria cases obtained from nearby local health facilities. We observed that the proportion of samples that gave rise to infection in the mosquitoes was higher for *P. vivax* (42–44%) when compared to *P. falciparum* (9–10%). Strikingly, samples with *P. vivax* and *P falciparum* gametocytes detectable by light microscopy where infectious in 58.8% and 8.7% of the DMFAs.

There are some well-supported hypotheses underpinning these observations. It has been shown that *P. vivax* gametocytes develop faster, and are present and infectious at the onset of an infection while *P. falciparum* gametocytes take around 10 days to mature (Bousema and Drakeley, 2011). Consequently, lower infectiousness in symptomatic *P. falciparum* patients as compared to *P. vivax* patients is expected since people are likely to seek treatment before

*P. falciparum* gametocytes have matured (Kiattibutr et al., 2017). Additional plausible explanations for the lower *P. falciparum* infectivity are that *P. falciparum* gametocytes may be more temperature sensitive and the time-delays between sample collection and feeding in this study may have increased the occurrence of exflagellation before feeding (Ogwan’g et al., 1993) or that the immune factors present in the mosquito blood meal could have prevented transmission of *P. falciparum* (Lensen et al., 1998). Further studies including investigations of immunological processes mediating transmission success are needed. As such, future studies will include DMFAs conducted in parallel with direct feeding experiments and serum replacement experiments with naïve donor serum.

In the present study, we observed that *P. vivax* samples from symptomatic patients were especially infectious to mosquitoes. Maximising the capability to diagnose *P. vivax* before sampling is thus expected to significantly increase the number of successful infections. The lack of a well-trained microscopists to read the slides before bleeding was a limitation in this study and light microscopy results were only obtained retrospectively. Based on our results we estimate that light microscopy diagnosis before bleeding would enable a further increase of DMFA success rate with *P. vivax* to around 60% if suitable *P. vivax* samples (those with gametocytes by light microscopy) were selected. Based on microscopy results, severity of mosquito infection is difficult to predict as the observed correlation between mosquito infection and oocyst density, and *P. vivax* gametocyte density was weak. For example, we observed cases where there were high mosquito infection rates resulting from low *P. vivax* gametocyte densities while in other cases low mosquito infection rates resulted from high *P. vivax* gametocyte densities (Figure 2A). This was noted in other studies as well, where it was observed that *P. vivax* gametocyte density was not a good indicator of mosquito infectivity. (Sattabongkot et al., 2003; Sattabongkot et al., 1991) We hypothesise that this is mainly attributable to the uncertainty associated with light microscopy quantification of gametocyte density.

We found that species determination by light microscopy was a very good predictor of infection success, as *P. vivax* infections resulted in approximately 10-fold increased infection success in the mosquitoes as compared to *P. falciparum*.

However, in a resource limited setting such as in the present study, light microscopy for clinical diagnosis is often not available. In the absence of light microscopy, it is important to assess which RDT result will most likely lead to a mosquito infection. As commonly known, RDT results are not reliable in distinguishing between *Plasmodium* species in co-endemic settings, however, the present study shows that they can be used to prioritise samples selected for DMFAs in order to maximise the probability of success (Table 2) (World Health Organization, 2006). We observed that in the group of samples positive for only pLDH with the CareStart RDT the proportion of successful DMFAs was highest (35.1%) as compared to HRP2 (27.3%) or when positive for both antigens (11.1%). This difference in proportions was statistically significant (p< 0.01). Therefore, by selecting samples only positive for pLDH over HRP2 (or both antigens) DMFA success can be increased up to 3 fold. In a setting like PNG where both *P. falciparum* and *P. vivax* are present in roughly equal proportions, it is likely to have a *P. vivax* infection when the RDT is positive for only the pLDH antigen, which is what we observed as well. (World Health Organization., 2017) Furthermore, it has been shown that HRP2 can remain positive between 35–42 days after treatment giving a false positive result while for pLDH it takes only 2 days before the antigen is cleared from circulation giving a more reliable result. (Grandesso et al., 2016)

Minimising sample transportation time was a likely determinant for DMFA success in this study. Only 2 out of 45 samples (4.4%) collected at Madang Town Clinic, which took 2 hrs before feeding, resulted in mosquito infection, whereas 36/137 (26.3%) of samples collected in the clinic adjacent to the laboratory, which took 15–25 minutes before feeding, resulted in mosquito infection. This difference in proportions is highly significant in a univariate analysis (p< 0.01), however, when adjusting for *P. vivax* in a logistic regression model the impact diminished.

This study provides important insights into the infectivity of symptomatic malaria cases to *An. farauti* in PNG. We have established a DMFA, which could serve as a platform to test potential transmission blocking vaccines and antimalarials, which act on gametocytes or the mosquito developmental stages of *Plasmodium vivax*.

## Data Availability

The data for the study is available for those interested

## Acknowledgements

This work was supported in part by the Bill and Melinda Gates Foundation (OPP1034577), National Institute of Allergy and Infectious Diseases (NIAID) (5U19AI089686–03), Swiss National Science Foundation (310030_134889), and National Health and Medical Research Council (NHMRC) of Australia (GNT1127356). LT is supported by a PhD scholarship from James Cook University, IM is supported by a Research Fellowship from NHMRC, LJR and SK are supported by Career Development Fellowships from NHMRC of Australia.We would like to sincerely thank all study participants. We thankfully acknowledge the assistance of clinical staff from Yagaum Hospital and Madang Town Clinic. We are grateful for technical assistance with setting up the DMFAs provided by Jetsumon Prachumsri and Kirakorn Kiattibutr from Mahidol University Vivax Research Unit in Thailand. Contribution by PNGIMR staff is thankfully acknowledged, especially that of research nurses Kaye Kose and Ruth Larry; laboratory technicians Hega Sakel. Lemen Kilepak, Muker Sakur, Yule E’ele, Siub Yabu and Wal Kuma, as well as expert microscopist Lina Lorry.

## References

Beebe, N.W., Cooper, R.D., Foley, D.H., Ellis, J.T., 2000. Populations of the south-west Pacific malaria vector Anopheles farauti s.s. revealed by ribosomal DNA transcribed spacer polymorphisms. Heredity (Edinb) 84 (Pt 2), 244–253.

Bousema, T., Dinglasan, R.R., Morlais, I., Gouagna, L.C., van Warmerdam, T., Awono-Ambene, P.H., Bonnet, S., Diallo, M., Coulibaly, M., Tchuinkam, T., Mulder, B., Targett, G., Drakeley, C., Sutherland,C., Robert, V., Doumbo, O., Touré Y., Graves, P.M., Roeffen, W., Sauerwein, R., Birkett, A., Locke, E., Morin, M., Wu, Y., Churcher, T.S., 2012. Mosquito feeding assays to determine the infectiousness of naturally infected Plasmodium falciparum gametocyte carriers. PloS one 7, e42821-e42821.

Bousema, T., Drakeley, C., 2011. Epidemiology and infectivity of Plasmodium falciparum and Plasmodium vivax gametocytes in relation to malaria control and elimination. Clinical microbiology reviews 24, 377–410.

Cattani, J.A., Tulloch, J.L., Vrbova, H., Jolley, D., Gibson, F.D., Moir, J.S., Heywood, P.F., Alpers, M.P., Stevenson, A., Clancy, R., 1986. The epidemiology of malaria in a population surrounding Madang, Papua New Guinea. Am J Trop Med Hyg 35, 3–15.

Churcher, T.S., Blagborough, A.M., Delves, M., Ramakrishnan, C., Kapulu, M.C., Williams, A.R., Biswas, S., Da, D.F., Cohuet, A., Sinden, R.E., 2012. Measuring the blockade of malaria transmission--an analysis of the Standard Membrane Feeding Assay. International journal for parasitology 42, 1037–1044.

Churcher, T.S., Trape, J.F., Cohuet, A., 2015. Human-to-mosquito transmission efficiency increases as malaria is controlled. Nat Commun 6, 6054.

Collins, W.E., Sullivan, J.S., Nace, D., Williams, T., Sullivan, J.J., Galland, G.G., Grady, K.K., Bounngaseng, A., 2002. Experimental infection of Anopheles farauti with different species of Plasmodium. J Parasitol 88, 295–298.

Delves, M., Plouffe, D., Scheurer, C., Meister, S., Wittlin, S., Winzeler, E.A., Sinden, R.E., Leroy, D., 2012. The activities of current antimalarial drugs on the life cycle stages of Plasmodium: a comparative study with human and rodent parasites. PLoS medicine 9, e1001169.

Diallo, M., Toure, A.M., Traore, S.F., Niare, O., Kassambara, L., Konare, A., Coulibaly, M., Bagayogo M., Beier, J.C., Sakai, R.K., Toure, Y.T., Doumbo, O.K., 2008. Evaluation and optimization of membrane feeding compared to direct feeding as an assay for infectivity. Malaria journal 7, 248.

Grandesso, F., Nabasumba, C., Nyehangane, D., Page, A.-L., Bastard, M., De Smet, M., Boum, Y., Etard, J.-F., 2016. Performance and time to become negative after treatment of three malaria rapid diagnostic tests in low and high malaria transmission settings. Malaria journal 15, 496–496.

Graves, P.M., Burkot, T.R., Carter, R., Cattani, J.A., Lagog, M., Parker, J., Brabin, B.J., Gibson, F.D., Bradley, D.J., Alpers, M.P., 1988. Measurement of malarial infectivity of human populations to mosquitoes in the Madang area, Papua, New Guinea. Parasitology 96 (Pt 2), 251–263.

Howes, R.E., Battle, K.E., Mendis, K.N., Smith, D.L., Cibulskis, R.E., Baird, J.K., Hay, S.I., 2016. Global Epidemiology of Plasmodium vivax. Am J Trop Med Hyg 95, 15–34.

Kiattibutr, K., Roobsoong, W., Sriwichai, P., Saeseu, T., Rachaphaew, N., Suansomjit, C., Buates, S., Obadia, T., Mueller, I., Cui, L., Nguitragool, W., Sattabongkot, J., 2017. Infectivity of symptomatic and asymptomatic Plasmodium vivax infections to a Southeast Asian vector, Anopheles dirus. International journal for parasitology 47, 163–170.

Lensen, A., Mulder, L., Tchuinkam, T., Willemsen, L., Eling, W., Sauerwein, R., 1998. Mechanisms that reduce transmission of Plasmodium falciparum malaria in semiimmune and nonimmune persons. The Journal of infectious diseases 177, 1358–1363.

Müller, I., Bockarie, M., Alpers, M., Smith, T., 2003. The epidemiology of malaria in Papua New Guinea. Trends in parasitology 19, 253–259.

Nace, D., Williams, T., Sullivan, J., Williams, A., Galland, G.G., Collins, W.E., 2004. Susceptibility of Anopheles farauti to infection with different species of Plasmodium. J Am Mosq Control Assoc 20, 272–276.

Ogwan’g, R.A., Mwangi, J.K., Githure, J., Were, J.B., Roberts, C.R., Martin, S.K., 1993. Factors affecting exflagellation of in vitro-cultivated Plasmodium falciparum gametocytes. The American journal of tropical medicine and hygiene 49, 25–29.

Ouedraogo, A.L., Guelbeogo, W.M., Cohuet, A., Morlais, I., King, J.G., Goncalves, B.P., Bastiaens, G.J.H., Vaanhold, M., Sattabongkot, J., Wu, Y., Coulibaly, B., Ibrahima, B., Jones, S., Morin, M., Drakeley, C., Dinglasan, R.R., Bousema, T., 2013. A protocol for membrane feeding assays to determine the infectiousness of P.falciparum naturally infected individuals to Anopheles gambiae. Malaria World Journal 4, 4.

Oueédraogo, A.L., Guelbeégo, W.M., Cohuet, A., Morlais, I., King, J.G., Goncçalves, B.P., Bastiaens, G.J.H., Vaanhold, M., Sattabongkot, J., Wu, Y., Coulibaly, M., Ibrahima, B., Jones, S., Morin, M., Drakeley, C., Dinglasan, R.R., Bousema, T., 2013. Methodology: A protocol for membrane feeding assays to determine the infectiousness of P. falciparum naturally infected individuals to Anopheles gambiae. Malaria World Journal 4, 7.

Roobsoong, W., Tharinjaroen, C.S., Rachaphaew, N., Chobson, P., Schofield, L., Cui, L., Adams, J.H., Sattabongkot, J., 2015. Improvement of culture conditions for long-term in vitro culture of Plasmodium vivax. Malaria Journal 14, 297.

Rutledge, L.C., Ward, R.A., Gould, D.J., 1964. Sutdies on the feeding response of mosquitoes to nutritive solution in a new membrane feeder. Mosquito News 24, 407–419.

Sattabongkot, J., Kumpitak, C., Kiattibutr, K., 2015. Membrane Feeding Assay to Determine the Infectiousness of Plasmodium vivax Gametocytes. Methods Mol Biol 1325, 93–99.

Sattabongkot, J., Maneechai, N., Phunkitchar, V., Eikarat, N., Khuntirat, B., Sirichaisinthop, J., Burge, R., Coleman, R.E., 2003. Comparison of artificial membrane feeding with direct skin feeding to estimate the infectiousness of Plasmodium vivax gametocyte carriers to mosquitoes. The American journal of tropical medicine and hygiene 69, 529–535.

Sattabongkot, J., Maneechai, N., Rosenberg, R., 1991. Plasmodium vivax: gametocyte infectivity of naturally infected Thai adults. Parasitology 102 Pt 1, 27–31.

Sauerwein, R.W., Bousema, T., 2015. Transmission blocking malaria vaccines: Assays and candidates in clinical development. Vaccine 33, 7476–7482.

Smith, R.C., Vega-Rodríguez, J., Jacobs-Lorena, M., 2014. The Plasmodium bottleneck: malaria parasite losses in the mosquito vector. Mem. Inst. Oswaldo Cruz 109, 644–661.

Sweeney, A.W., 1987. Larval salinity tolerances of the sibling species of Anopheles farauti. J Am Mosq Control Assoc 3, 589–592.

Vallejo, A.F., Rubiano, K., Amado, A., Krystosik, A.R., Herrera, S., Arevalo-Herrera, M., 2016. Optimization of a Membrane Feeding Assay for Plasmodium vivax Infection in Anopheles albimanus. PLoS neglected tropical diseases 10, e0004807.

Wampfler, R., Mwingira, F., Javati, S., Robinson, L., Betuela, I., Siba, P., Beck, H.-P., Mueller, I., Felger,I., 2013. Strategies for detection of Plasmodium species gametocytes. PloS one 8, e76316-e76316.

World Health Organization, 2006. The Use of Malaria Rapid Diagnostic Tests, 2 ed, Geneva, Switzerland, p. 20.

World Health Organization, 2019. World Malaria Report 2019. WHO, Geneva, p. 185.

World Health Organization., 2017. World Malaria Report 2017. WHO, Geneva, p. 196.

